# Long-Term Carotid Plaque Progression and the Role of Intraplaque Hemorrhage: A Deep Learning-Based Analysis of Longitudinal Vessel Wall Imaging

**DOI:** 10.1101/2024.12.09.24318661

**Authors:** Yin Guo, Ebru Yaman Akcicek, Daniel S. Hippe, SeyyedKazem HashemizadehKolowri, Xin Wang, Halit Akcicek, Gador Canton, Niranjan Balu, Duygu Baylam Geleri, Taewon Kim, Dean Shibata, Kaiyu Zhang, Beibei Sun, Xiaodong Ma, Marina S. Ferguson, Mahmud Mossa-Basha, Thomas S. Hatsukami, Chun Yuan

**Author notes:** **Address for correspondence:** Chun Yuan, PhD, 850 Republican St, Rm 124, Seattle, WA 98109.

## Abstract

**Background:** Carotid atherosclerosis is a major contributor in the etiology of ischemic stroke. Although intraplaque hemorrhage (IPH) is known to increase stroke risk and plaque burden, its long-term effects on plaque dynamics remain unclear. This study aimed to evaluate the long-term impact of IPH on carotid plaque burden progression using deep learning-based segmentation on multi-contrast magnetic resonance vessel wall imaging (VWI).

**Methods:** Twenty-eight asymptomatic subjects with carotid atherosclerosis underwent an average of 4.7 ± 0.6 VWI scans over 5.8 ± 1.1 years. Deep learning pipelines were used to segment the carotid vessel walls and IPH. Bilateral plaque progression was analyzed using correlation coefficients and generalized estimating equations. Associations between IPH occurrence, IPH volume, and plaque burden (%WV) progression were evaluated using linear mixed-effect models.

**Results:** IPH was detected in 23/50 of arteries at any time point. Of arteries without IPH at baseline, 11/39 developed new IPH that persisted, while 5/11 arteries with baseline IPH exhibited it throughout the study. Bilateral plaque growth was significantly correlated (r = 0.54, p < 0.001), but this symmetry was weakened in cases with IPH (r = 0.1, p = 0.62). Moreover, IPH presence or development at any point was associated with a 2.3% absolute increase in %WV on average within the affected artery (p < 0.001). The volume of IPH was also positively associated with increased %WV (p = 0.005).

**Conclusions:** Deep learning-based segmentation pipelines were utilized to identify IPH, quantify IPH volume, and measure their effects on carotid plaque burden during long-term follow-up. Findings demonstrated that IPH may persist for extended periods. While arteries without IPH demonstrated minimal progression under contemporary treatment, presence of IPH and greater IPH volume significantly accelerated long-term plaque growth.

## Introduction

Stroke remains the fifth leading cause of mortality in the United States, with an 8.4% increase in the age-adjusted death rate from 2011 to 2021^1^. Carotid atherosclerosis is a leading etiology in stroke^2^. The progression of carotid atheroma, which leads to a reduction in vessel lumen and eventual plaque rupture, is closely related to the risk of cerebrovascular ischemic events^3,4^. Understanding the mechanisms driving plaque evolution is essential for improving stroke prevention strategies.

A recent meta-analysis demonstrated the prevalence of intraplaque hemorrhage (IPH) in patients with both symptomatic and asymptomatic carotid stenosis and identified IPH as a stronger predictor of stroke than any known clinical risk factors^5^. Moreover, IPH is associated with increasing lumen stenosis and rapid vessel wall growth^6–9^. Despite this, little is known about the long-term behavior of IPH, including its occurrence, resolution, and distribution over extended periods, as most studies have been limited to follow-ups of one to two years^7–9^. Its long-term impact on plaque progression also remains inadequately explored.

Carotid magnetic resonance (MR) vessel wall imaging (VWI) is a noninvasive imaging technique validated with histology for the accurate in vivo detection of IPH and has become an established approach for measuring IPH and plaque growth in longitudinal studies^10–12^. Traditionally, manual analysis of longitudinal VWI has limited the ability to study carotid atherosclerosis due to its labor-intensive and expertise-dependent nature. Advances in deep learning-based segmentation have now enabled automated, reproducible measurements of the carotid vessel wall^13–15^ and IPH, facilitating comprehensive studies of plaque dynamics over longer follow-up periods.

This study leverages these tools to analyze carotid atherosclerosis in an initially asymptomatic cohort over approximately six years. By investigating both systemic and localized trends in plaque evolution, this work provides novel insights into the natural history of carotid atherosclerosis, emphasizing the critical role of IPH in driving plaque burden progression and its implications for stroke prevention.

## Methods

### Study population

Participants for this study were recruited as part of a prospective cohort study over a ten-year period (2012-2022), aimed at examining the natural history of carotid atherosclerosis through serial VWI. Eligible participants included patients aged 35 years and older with clinically identified atherosclerosis in at least one carotid artery and without cerebrovascular events in the six months prior to enrollment. Study procedures and consent forms were reviewed and approved by the institutional review board, and all participants provided written informed consent prior to enrollment. Detailed information on cohort recruitment has previously been published^16^.

To monitor the evolution of atherosclerotic plaque, serial VWI scans with consistent parameters (detailed below) were conducted up to five times during the study period. For the analysis of long-term plaque burden progression and the effects of IPH, we conducted a retrospective analysis of a subset of participants. Inclusion criteria for this retrospective analysis required subjects who had undergone at least three VWIs over a minimum span of five years. Arteries with a history of carotid endarterectomy (CEA) or stenting at study entry were excluded. Scans of arteries performed after CEA or stenting were also excluded.

### MR Imaging Protocol

Two large-coverage three-dimensional (3D) isotropic VWI sequences were performed using a 3.0T Philips Ingenia CX scanner (Philips Healthcare, Best, The Netherlands): 1) Motion-sensitized driven equilibrium prepared rapid gradient echo sequence (MERGE^17^) was used to delineate vessel wall morphology (TR: 10.0ms, TE: 4.0ms, flip angle: 6°, FOV: 250×42×250 mm); 2) an inversion-recovery gradient echo sequence with phase-sensitive reconstruction (simultaneous non-contrast angiography and intraplaque hemorrhage imaging, SNAP^18^) was used to detect and measure IPH (TR: 10,0ms, TE: 4.8ms, TI 500ms, IRTR 2000ms, flip angles: 11° and 5°, FOV: 160×32×160 mm). Both sequences were acquired in the coronal plane with an acquired resolution of 0.80×0.80×0.80 mm³, zero-interpolated to 0.40×0.40×0.40 mm³, and scan times of 5.3 minutes each. Detailed description of both sequences can be found elsewhere^9,16,19^.

### Ischemic infarct assessment

Conventional brain MRI examinations, including diffusion weighted imaging (DWI), susceptibility weighted imaging (SWI), and T2-fluid attenuated inversion recovery (FLAIR), were performed at all VWI time points. A board-certified neuroradiologist directly compared the baseline and follow-up brain MRI images and scored for any incident infarcts (FLAIR and DWI images) or hemorrhages (SWI images). Vascular territory and signs of acuity or associated hemorrhage were noted for the infarcts.

### Deep Learning-based Vessel Wall and IPH segmentation

Two deep learning-based imaging analysis pipelines were developed to bilaterally segment the carotid vessel wall and IPH on MERGE and SNAP sequences, respectively. Each scan was segmented independently. The complete workflow is illustrated in Supplementary Figure 1.

For the segmentation of carotid lumen and outer wall boundary using MERGE, images were first resampled to isotropic voxels with 0.35mm resolution. The location of the left and right common carotid bifurcations was then identified automatically using a 3D SonoNet^20^. On each side, a standardized 3D bounding box was extracted, encompassing 85 slices (29.8mm) superior and 170 slices (59.5mm) inferior to the bifurcation, with the axial plane covering the entire image dimensions. The bounding box was then used for a preliminary 3D lumen segmentation. A trained radiologist reviewed the preliminary segmentation to assess image quality and ensure that the lumen from all slices encompassing the bifurcation region were correctly localized. Finally, 2D patches of 80×80 pixels (28×28 mm) centered at the lumen along both carotids were segmented slice-by-slice using a 2.5D convolutional neural network utilizing 11-slice (3.9mm) volumetric context.

To segment IPH from SNAP, a same bifurcation detection module was first applied to locate the left and right carotid bifurcations. For each side, slices spanning the same anatomical extent as in the MERGE processing (29.8mm superior and 59.5mm inferior to the bifurcation) were selected and reformatted to 2mm axial spacing while maintaining the original in-plan resolution of 0.4× 0.4 mm, resulting in approximately 15 superior and 30 inferior slices. A 2D nnUNet^21^ was then trained to segment IPH slice-by-slice by identifying hyperintense signals. For each side, the axial patch size encompassed half the left-right dimension of the image and the entire anterior-posterior dimension.

For each MERGE image, after obtaining lumen and outer wall segmentations of the same artery, the range of slices with wall thickness greater than 2mm in any scan during the follow-up period was included as atherosclerotic plaque, ensuring consistent anatomical coverage across all time points relative to the bifurcation. This threshold of 2 mm aligns with the criteria used in the Rotterdam^22^, PARISK^23^ and other established researches. The primary outcome of plaque burden progression was the percent wall volume (%WV), calculated as wall volume divided by total vessel volume, multiplied by 100. Additionally, as secondary outcomes to further assess the mechanisms of plaque progression, mean lumen area and mean total vessel area (lumen + wall area) were calculated by averaging the respective areas across all slices within the plaque range, providing measures of luminal narrowing and vessel remodeling. For descriptive characterization, the same metrics were also calculated at baseline for adjacent vessel segments without established plaque, separately for the common carotid artery (inferior to the plaque) and internal carotid artery (superior to the plaque), to provide reference values of healthy vessel morphology.

For each SNAP image per artery per time point, IPH detection was confined to the plaque range identified from MERGE using the bifurcation as a landmark for alignment, with the corresponding slice range in SNAP determined by scaling the relative slice positions to the bifurcation by the ratio of slice spacings (0.35mm / 2mm). Furthermore, a trained radiologist reviewed all segmented IPH regions by visually comparing them with the lumen on SNAP images, excluding IPH regions that were clearly distant from the vessel and anatomically implausible. Although such excluded regions were rare, this quality control step ensured that IPH measurements reflected hemorrhage within the artery. Subsequently, IPH was categorized as present (IPH+) or absent (IPH-). IPH volume was aggregated as the sum of IPH areas per slice multiplied by 2mm, and percent hemorrhage volume (%HV) was defined as IPH volume divided by wall volume, multiplied by 100.

Detailed information about the training dataset and extensive validation studies can be found in the Supplementary Materials. External validation datasets were utilized (see Figure 1 for the cohort information). Histological validation in 14 subjects (121 SNAP slices, 49 with IPH) showed moderate agreement for IPH detection (kappa: 0.62, 95% CI: 0.48-0.76), with 61% sensitivity and 97% specificity. Scan-rescan reproducibility in 33 subjects demonstrated high ICCs of 0.88 (95% CI: 0.80-0.93) for %WV and 0.94 (95% CI: 0.84-0.98) for IPH volume. Full validation details are provided in the Supplementary Materials.

**Figure 1.**
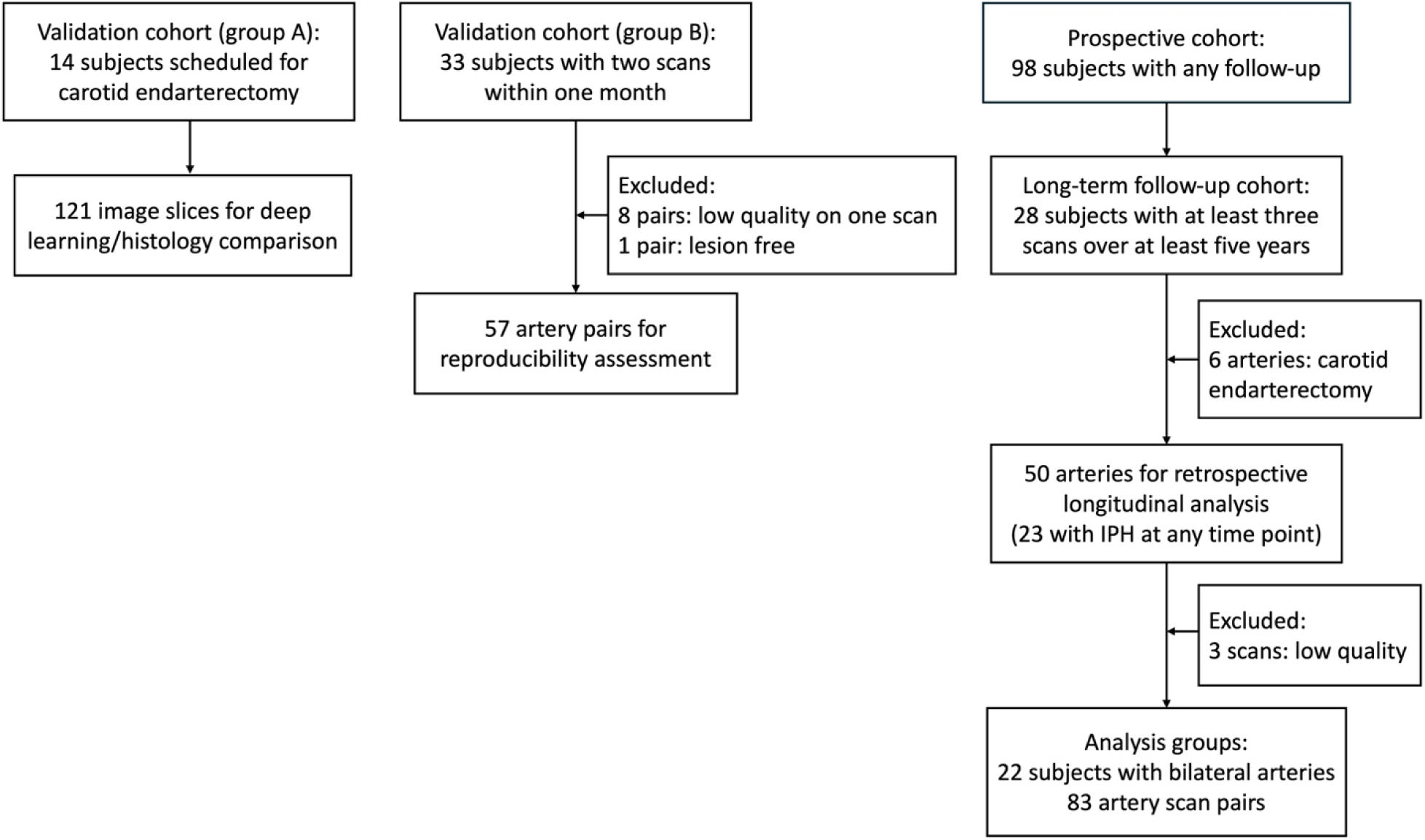
Flow chart of the study cohort.

### Statistical Analysis

Summary statistics are presented as mean ± standard deviations (SDs) or median (inter-quartile range [IQR]) for continuous variables and counts with percentage for categorical variables.

For each analyzable plaque, we calculated two types of plaque growth rate: 1) Long-term growth rate was defined as the annualized absolute change in %WV over the entire follow-up period (expressed in units of percent per year), determined by fitting a linear model across all available time points^24^; 2) Short-term growth rates were calculated as the annualized difference in %WV between consecutive scans. The method for deriving long-term and short-term plaque growth rates is illustrated in Figure 2. Subsequently, generalized estimating equation (GEE)-based linear regression models were used to evaluate associations of both long-term and short-term growth rate of each side with the contralateral side, adjusted for potential confounding factors of age, sex, history of hypertension and statin use. The number of risk factors considered in the models was limited by the available sample size. Effect sizes were summarized using the partial correlation coefficient.

**Figure 2.**
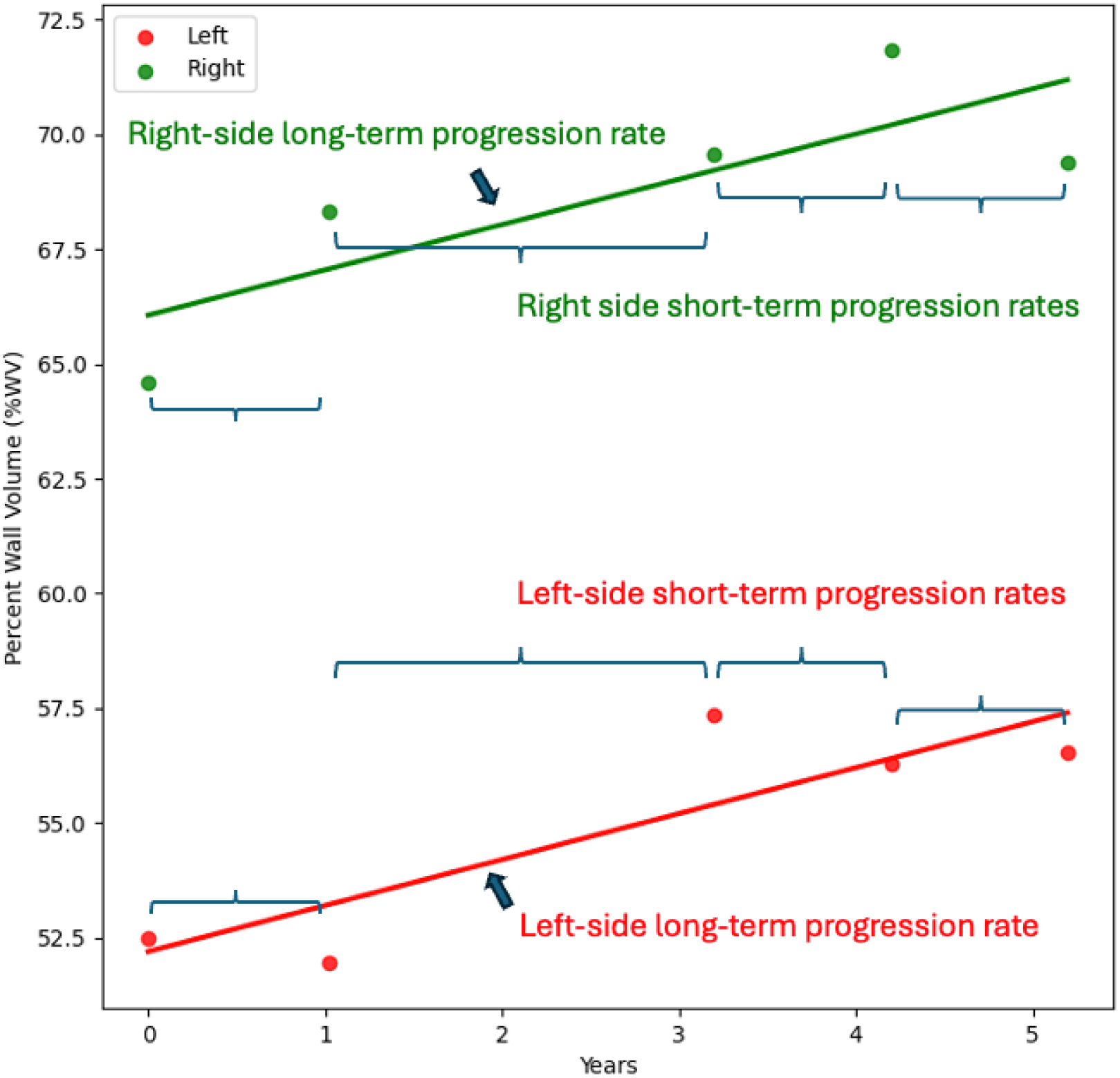
Illustration of the calculations for long-term and short-term progression rates of plaque burden. For each subject, the percent wall volume (%WV) of plaques in the left and right carotid arteries was measured at all available time points. For each plaque, a linear model was fitted to determine the long-term progression rate, expressed as the annualized absolute change in %WV (%/year). Short-term progression rates (%/year) were calculated as the annualized difference in %WV between consecutive scans.

To investigate whether progression rate in plaque burden depends on presence and volume of IPH in each artery, we employed a linear mixed-effects model. This model used %WV as the primary outcome variable and included random intercepts to account for within-subject and within-artery correlation. As fixed effects, the model included time as a continuous variable to estimate the growth rate of %WV and account for variation in scan intervals, a binary predictor indicating IPH presence, and %HV to assess the effect of IPH volume. %HV was log2-transformed to reduce right skewness, and further transformed such that it equals zero when IPH is absent, and equals the deviation from the population mean when IPH is present. The model setup was designed to disentangle the effects of the presence and volume of IPH on plaque burden progression. This model was further adjusted for age, sex, history of hypertension and statin use. Moreover, the same mixed-effects model framework was applied using mean lumen area and mean total vessel area as outcome variables to assess the effects of IPH on luminal narrowing and wall expansion. The Wald method was used to compute 95% confidence intervals (CI) for the fixed-effect coefficients.

All statistical analyses were performed using R 4.4.0. A two-sided p-value < 0.05 was considered statistically significant.

## Results

### Patient Characteristics

A total of 98 subjects were recruited prospectively, and population demographics can be found in the referenced publication^9^. Among these, 28 subjects underwent at least three VWIs over a span of at least five years and were therefore included in this retrospective analysis. All subjects were under contemporary medical management, and the baseline clinical information of these 28 subjects is presented in Table 1. After three scans were excluded due to poor image quality, each subject had an average of 4.7 ± 0.6 scans within 5.8 ± 1.1 years, with an average interval of 1.6 ± 0.9 years between consecutive scans, indicating a relatively uniform distribution of imaging time points and follow-up duration across the cohort. Figure 1 shows the study flow chart.

**Table 1.** Population demographics at baseline.

| Variable | Value |
| --- | --- |
| Age, years | 71.3 ± 9.7 |
| Male | 20 (71%) |
| Current smoker | 3 (10%) |
| History of hypertension | 17 (60%) |
| History of hyperlipidemia | 25 (89%) |
| History of diabetes | 3 (10%) |
| Statins used | 21 (75%) |
| Antihypertensives | 16 (57%) |
| Antiplatelets | 21 (75%) |

Values are no. (%) or mean ± standard deviation

Among the 56 carotid arteries within the 28 subjects, six were excluded due to prior history of CEA or having fewer than three available scans due to CEA during the study period. The remaining 50 arteries from 28 subjects were included in artery-level analyses. Of these subjects, there were 22 with bilateral arteries available, enabling comparison of plaque growth between sides. This provided a sample size of 22 subjects for the comparison of long-term progression rates between sides and a sample size of 83 scan-pairs from these 22 subjects for the comparison of short-term progression rates between sides.

### Baseline Plaque and Vessel Characteristics

Among the 50 analyzable carotid arteries, atherosclerotic plaques (wall thickness >2mm) had a median length of 26.6 mm (IQR: 27 mm). At baseline, the median %WV was 53.6% (IQR: 7.6%), median mean wall thickness was 2.57 mm (IQR: 0.7 mm), median mean lumen area was 40.9 mm² (IQR: 14.7 mm²), and median mean total vessel area was 89 mm² (IQR: 21.4 mm²).

For reference, baseline measurements in adjacent vessel segments without established plaque (wall thickness <2mm) were as follows: In the common carotid artery (CCA, inferior to plaque), the median %WV was 47.0% (IQR: 6.1%), median mean wall thickness was 1.41 mm (IQR: 0.27 mm), median mean lumen area was 32.7 mm² (IQR: 12.6 mm²), and median mean total vessel area was 62.5 mm² (IQR: 20.3 mm²). In the internal carotid artery (ICA, superior to plaque), the median %WV was 48.4% (IQR: 6.7%), median mean wall thickness was 1.33 mm (IQR: 0.23 mm), median mean lumen area was 23.7 mm² (IQR: 12.1 mm²), and median mean total vessel area was 47.8 mm² (IQR: 18.7 mm²).

### IPH Presence, Occurrences, and Relation to New Infarcts

Among the 50 carotid arteries from 28 subjects, 23 arteries (46%) from 17 subjects (60.7%) showed presence of IPH on VWI at any time point during the study period. Of the 39 IPH-arteries at the baseline scan, eleven (28.2%) became IPH+ on all subsequent scans after the initial occurrence of IPH, and one artery (2.56%) developed new IPH on the second scan but reverted to IPH-on scans 3 to 5. Of the 11 arteries that were IPH+ at baseline, IPH persisted in five (45.5%) throughout the study, whereas six (54.5%) demonstrated resolution of IPH before the final scan.

During the follow-up, only three subjects (10.7%) developed new ischemic infarcts, with two (66.7%) occurring among the 17 subjects (60.7%) with either unilateral or bilateral IPH. One of these subjects consistently exhibited ipsilateral IPH and was IPH-on the contralateral side across all scans. In the other subject, a new infarct was detected at the third scan, coinciding with the appearance of new ipsilateral IPH. The third subject who developed an infarct had no detectable IPH in either carotid artery at any time point. Given the limited number of ischemic events, the association between IPH and ischemic stroke in this study remains exploratory in nature.

### Bilateral Symmetry in Plaque Evolution and the Disruptive Role of IPH

In the 22 subjects with bilateral analyzable arteries, the median baseline %WV was 54.2% (IQR: 3.7%) for the left carotid artery and 50.9% (IQR: 9.1%) for the right. The median long-term %WV absolute growth rate was −0.3%/year (IQR: 1.01%/year) on the left side and - 0.02%/year (IQR: 0.75%/year) on the right, while the median short-term growth rate was - 0.47%/year (IQR: 2.53%/year) on the left and 0.08%/year (IQR: 3.84%/year) on the right. Both long-term and short-term plaque growth rates of one side were correlated with the contralateral side (long-term rates: r = 0.54, p < 0.001; short-term rates: r = 0.28, p = 0.004), suggesting that plaque progression may be influenced by systemic factors. The significant correlation remained after adjusting for age, sex, history of hypertension and statin use (r = 0.38, p = 0.016; and r = 0.26, p = 0.007). Figure 3 provides an example of bilateral plaque burden increases over five scans. The association between plaque progression on the left and right sides is visualized in Figure 4.

**Figure 3.**
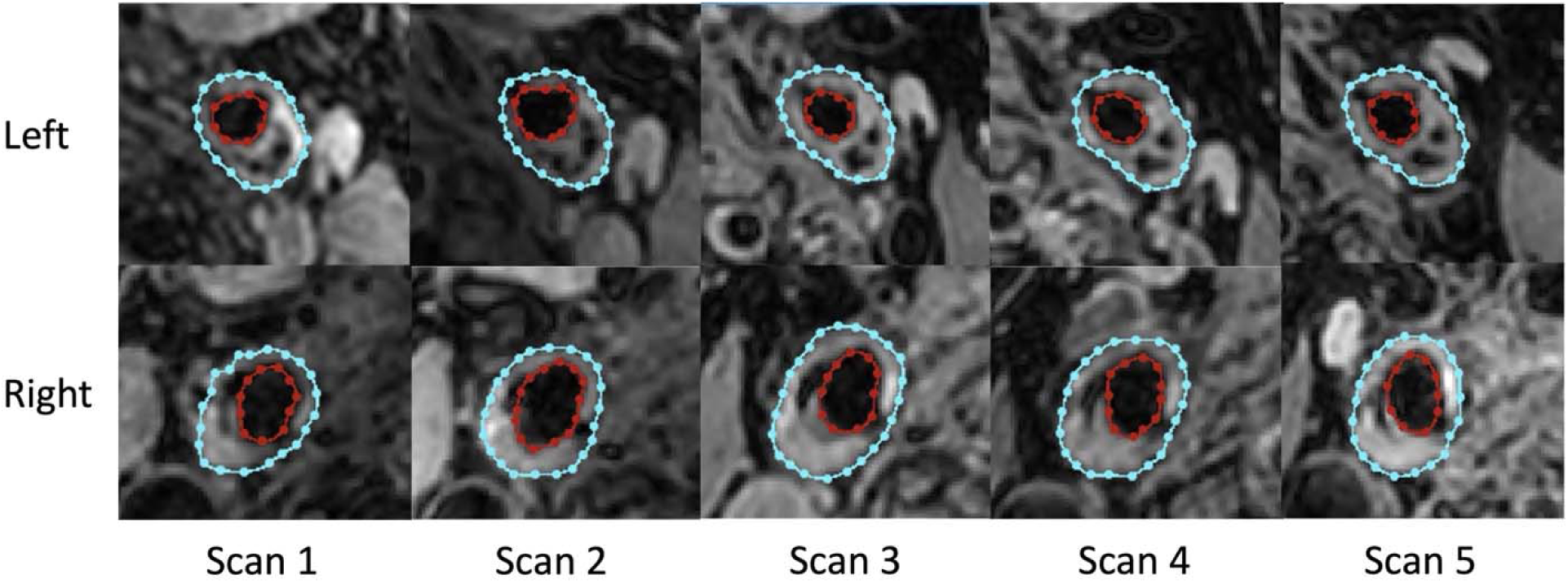
Example of bilateral plaque burden increases over five vessel wall imaging sessions spanning five years, demonstrating symmetrical plaque progression on both the left and right sides. The vessel wall contours were generated using deep learning-based segmentation, with red contours delineating the vessel lumen and blue contours delineating the vessel outer wall boundary.

**Figure 4.**
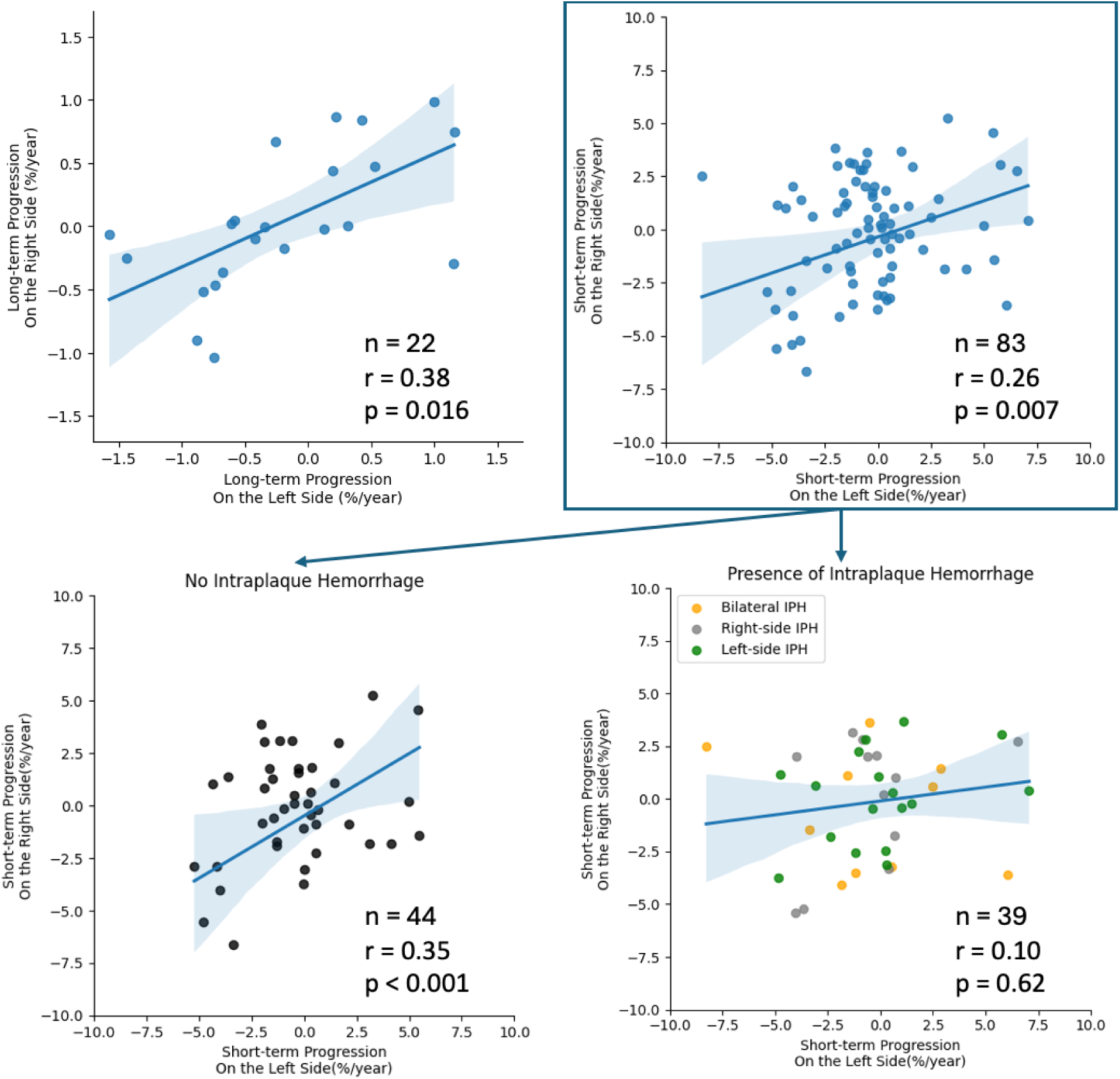
Upper panels: Correlation of long-term and short-term growth rate of plaque burden between bilateral carotid arteries, demonstrating systemic influences on bilateral plaque progression. Lower panels: Correlation of bilateral short-term progression in the two subgroups of carotid arteries with (right) and without IPH (left). The partial correlation coefficient between left and right plaque progression rates was calculated after adjusting for age, sex, history of hypertension, and statin use.

To explore the potential effects of IPH, the short-term progression rates of the arteries were further analyzed and divided into two sub-groups based on detected IPH presence. In 44 scan-pairs, IPH was not detected in either of the two consecutive scans. In this IPH-subgroup, the correlation of plaque progression on one side with the contralateral side was significant (r = 0.35, p < 0.001). However, among the 39 scan-pairs with either unilateral or bilateral IPH detected, the correlation of progression rates between sides decreased and was no longer remained significant (r = 0.10, p = 0.62). These findings suggest that the presence of IPH may be associated with a disruption of the systemic symmetry in bilateral plaque evolution. The correlation of bilateral short-term plaque progression in the two subgroups is visualized in Figure 4.

### Impact of IPH Presence and Volume on Plaque Progression

Building on these findings, we further evaluated both the presence and the volume of IPH on %WV progression of each artery using a linear mixed-effects model. In the unadjusted model, the random effects for patients and arteries accounted for individual variability, with SDs of 4.3% and 1.9%, respectively. The SD of the residual term is 3.1%. This suggests considerable variability in %WV progression among patients and between bilateral arteries within the same patient.

The progression of %WV over time in arteries without IPH was minimal, with an average absolute growth rate of −0.016 %/year (95% CI: −-0.19 - 0.16, p = 0.86). However, the presence and occurrence of IPH at any point was a significant predictor, associated with a 2.32% absolute increase in %WV on average (95% CI: 1.00 - 3.65, p < 0.001). This indicates that patients with IPH had higher %WV compared to those without IPH and that the development of new IPH was significantly associated with an increase in plaque burden.

Furthermore, within the IPH+ arteries, %HV was further associated with %WV (p = 0.007). Each doubling of the %HV – reflected by a 1-unit increase in the log2-transformed %HV – was associated with an additional 0.69% absolute increase in %WV (95% CI: 0.19 - 1.18), corresponding to a 29% increase relative to the average IPH effect ([2.34% + 0.69%] / 2.34% = 1.29). This underscored the significant impact of both the presence and volume of IPH on the progression of carotid plaque burden over time, as illustrated in Figure 5. Figure 6 provides an example of plaque progression over four scans, highlighting the increasing plaque burden alongside the occurrence (scan 3) and subsequent volume expansion (scan 4) of IPH.

**Figure 5.**
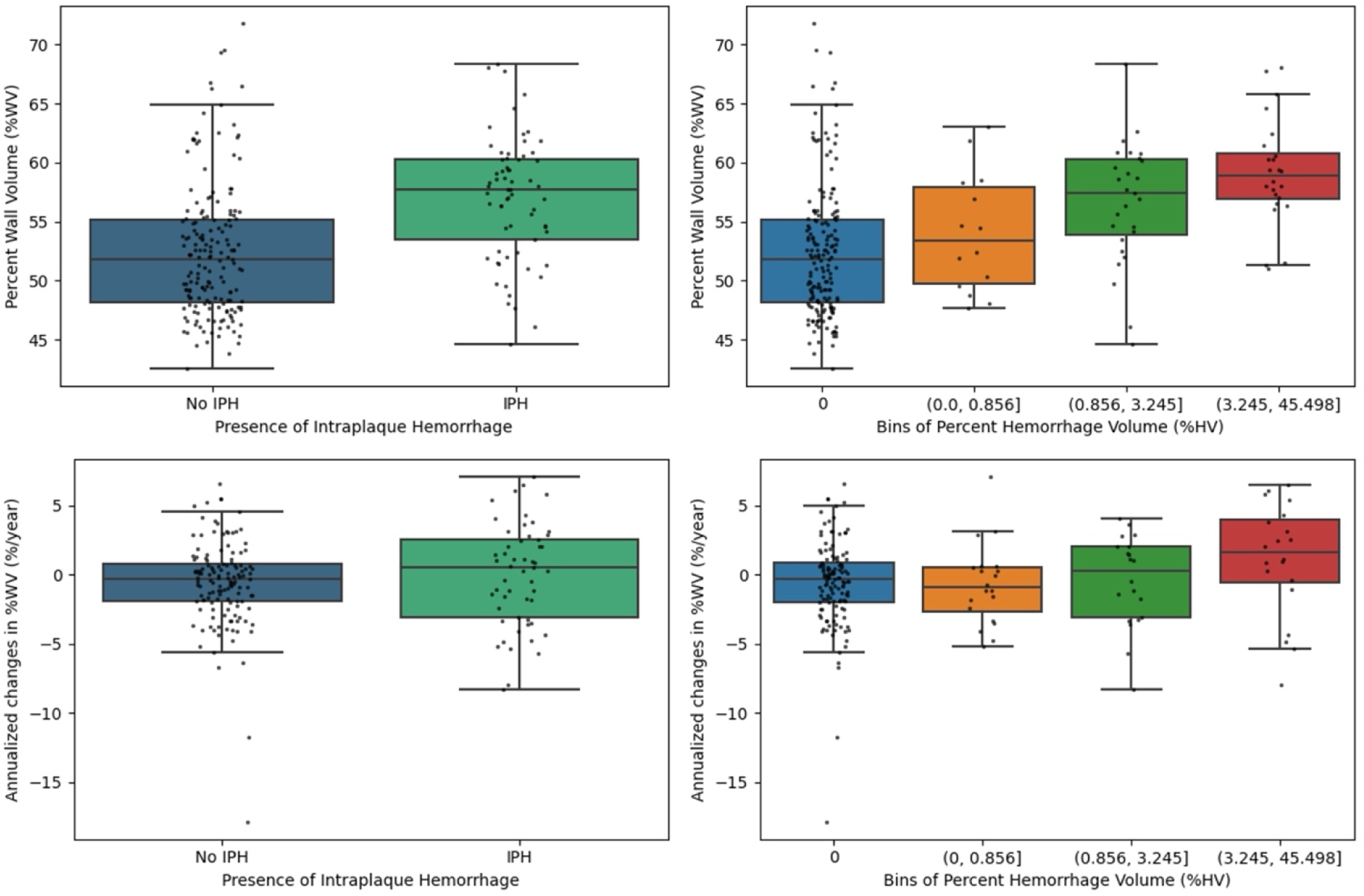
Box plots demonstrating the effects of presence and volume of IPH on plaque burden. In (a) and (c), %WV and annualized changes in %WV were compared based on presence or not of IPH. In (b) and (d), %HV was divided into three equal-sized bins, comparing %WV and annualized changes in %WV across different degrees of IPH volume.

**Figure 6.**
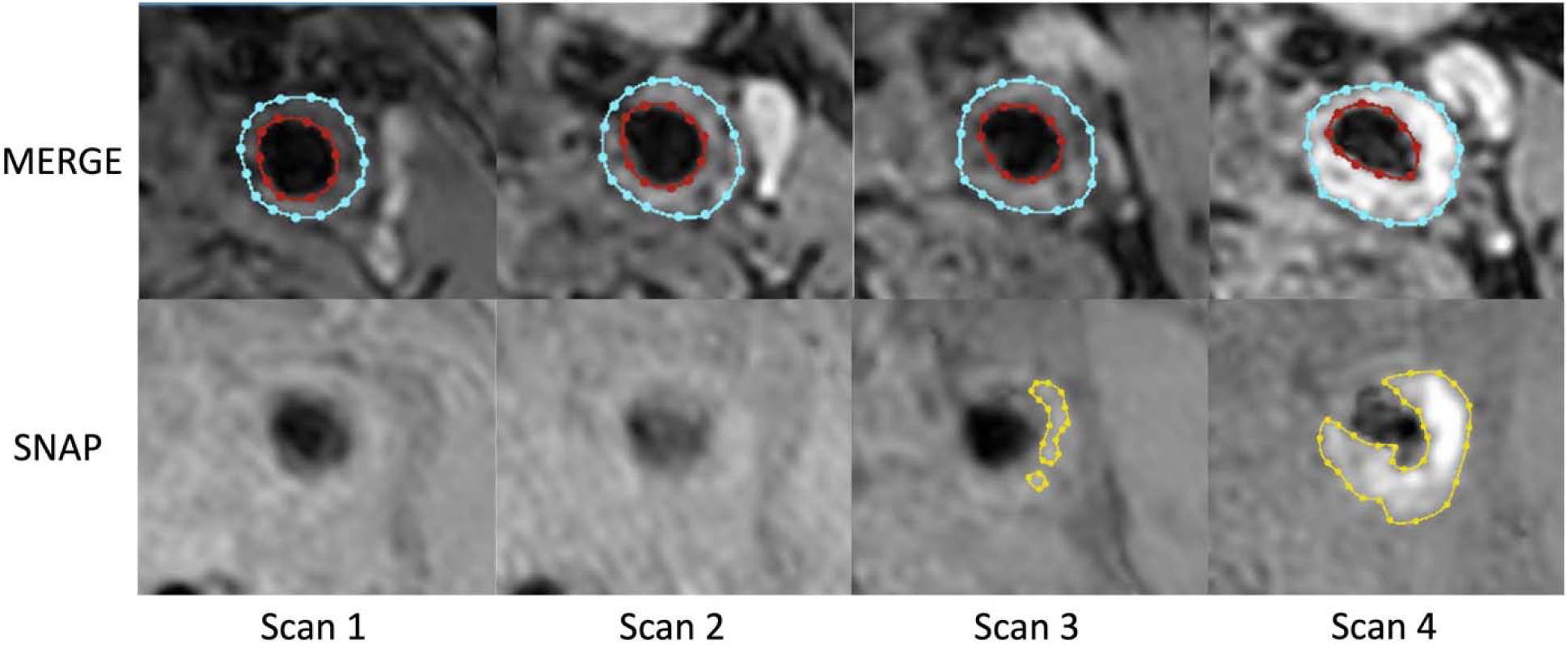
Example of plaque progression over four vessel wall imaging sessions spanning five years, demonstrating an increase in plaque burden, marked by the occurrence of intraplaque hemorrhage (IPH) at Scan 3 and subsequent volume expansion at Scan 4. The vessel wall and IPH contours were generated using deep learning-based segmentation, with red and blue contours delineating the vessel lumen and vessel outer wall boundary on MERGE, and orange contours delineating IPH boundary on SNAP.

When adjusted for age, sex, history of hypertension and statin use, the linear mixed-effects model continued to show strong associations between IPH and %WV progression. The presence of IPH was associated with an adjusted 2.28% absolute increase in %WV (95% CI: 0.93 - 3.62, p = 0.001), and the %HV association persisted, with a coefficient of 0.69% (95% CI: 0.20 - 1.20, p = 0.007), similar to the unadjusted model. Age, sex, history of hypertension and statin use did not have statistically significant associations with %WV progression (p > 0.34 for each).

### Mechanisms of IPH-Associated Plaque Progression

Using mean lumen area and mean total vessel area as secondary outcomes in the adjusted mixed-effects model, the presence of IPH was associated with a significant decrease in lumen area of 1.57 mm^2^ (95% CI: 0.14 - 3, p = 0.033), and greater %HV was further associated with additional lumen narrowing, with each doubling of %HV corresponding to a 0.6 mm^2^ decrease in lumen area (95% CI: 0.079 - 1.11, p = 0.025). In contrast, neither IPH presence (p = 0.66) nor %HV (−0.29) were significantly associated with changes in mean total vessel area. Male sex was associated with larger baseline total vessel area (15.6 mm^2^, 95% CI: 1.97 - 29.23, p = 0.035).

## Discussion

By leveraging two deep learning-based segmentation pipelines on two large-coverage 3D MR vessel wall imaging (VWI) sequences, we comprehensively evaluated carotid plaque features across an average of five scans over six years in an asymptomatic patient cohort. The role of intraplaque hemorrhage (IPH) in long-term plaque progression can be characterized in two critical ways. First, while plaque progression on one side was significantly correlated with the contralateral side, reflecting the systemic nature of atherosclerotic disease, this symmetry was notably weakened in subjects with IPH, suggesting an association between IPH and more localized effects. Second, despite contemporary medical management, the occurrence and volume of IPH were strongly associated with plaque burden increase, whereas plaques without IPH remained stable over the study period. Furthermore, our secondary analyses revealed that IPH-associated plaque progression manifests primarily as progressive luminal narrowing, with no significant association with changes in total vessel area. These findings deepen our understanding of the long-term and pivotal role IPH plays in the natural history of carotid atherosclerosis and underscore the importance of early IPH detection for optimized strategies in managing atherosclerosis.

Previous studies have demonstrated carotid plaque symmetry by comparing cross-sectional measurements of plaque burden between bilateral carotid arteries, as observed in ex vivo imaging studies^25^ and in vivo assessments using ultrasound^26^ and VWI^27^. Our study extends this understanding with longitudinal follow-up over a six-year period, which reveals not just static morphological symmetry but also symmetrical trends in bilateral plaque progression. While our results suggest that monitoring the more advanced plaque could provide prognostic insights into the less advanced contralateral plaque, the clinical implications of bilateral plaque progression remain an area for further investigation. Specifically, it is unclear whether patients with plaque disruption in one carotid artery are more likely to develop similar disruptions in the contralateral artery or other vascular beds. Evidence from prior studies, such as the AIM-HIGH MRI sub-study^28^, suggests that patients with high-risk carotid plaque features may have an increased risk of systemic cardiovascular events, highlighting the need for further research into the relationship between local plaque progression and broader vascular instability.

Notably, as demonstrated by Li et al.^27^, IPH exhibits low concordance between left and right carotid plaques, with unilateral or bilateral IPH frequently reported in the current and other studies, underscoring the role of localized factors in its pathogenesis. Studies showed that IPH development, in part driven by the rupture of neo-vessels, is shaped by conditions such as hemodynamic forces^29^, local mechanical deformation^30^, calcification^9^, and inflammation^31^ within the plaque microenvironment. At the same time, systemic factors still significantly contribute to the development of IPH. For instance, the Rotterdam Study^32^, in a cross-sectional analysis, identified pulse pressure as the most significant predictor of IPH, independent of plaque burden and other cardiovascular risk factors. Similarly, another longitudinal study associated male sex and hypertension with an increase in IPH^9^. Understanding the interplay between these localized and systemic influences is critical for advancing our understanding of pathophysiology of IPH development and its impact on plaque stability.

IPH is a dynamic process with the potential for growth or resolution over time. Longitudinal follow-up studies have shown that its hyperintense signal on VWI, likely attributable to methemoglobin from hemoglobin breakdown after erythrocyte extravasation, can persist throughout the course of atherosclerotic disease. Previous studies^7,33,34^ using T1-weighted sequences reported IPH resolution rates of 0%, 0%, and 6% over 17 to 18 months of follow-up. More recently, one study^9^ found that IPH resolved in only 3 out of 89 scan pairs (3.4%) over an average of 1.8 years, utilizing the SNAP sequence which combines inversion-recovery gradient echo with phase-sensitive acquisition to enhance IPH-to-wall contrast and measurement accuracy^35,36^. Similarly, our study, using SNAP to detect IPH, demonstrated that all but one newly developed IPH persisted to the study end, around half arteries with pre-existing IPH continued to exhibit it throughout the study period, and hyperintense IPH signals may be observed for as long as six years. This persistence may be explained by ongoing erythrocyte extravasation from leaky neo-vessels^37^.

IPH is widely recognized as a strong indicator of carotid plaque vulnerability, with its presence linked to an increased risk of future ischemic cerebrovascular events^38^, independent of symptom status^39^ or degree of stenosis^5,40^. In our study, despite more than half of subjects exhibiting IPH at some point during follow-up, the incidence of ischemic symptoms over at least five years was low. Histological studies^37,41^ have supported this observation by identifying neovasculature, a potential source of IPH, predominantly located in the adventitia away from the plaque surface, thus preserving surface integrity. This suggests that assessing plaque surface disruption^42,43^ could be an important additional metric for stratifying stroke risk, particularly in asymptomatic patients. Nonetheless, IPH has been consistently associated with a rapid increase in plaque burden^7,9^ and decrease in lumen area, and our findings added to the body of evidence by further highlighting the sustained and long-term impact of IPH presence and volume on the progression of atherosclerotic disease.

Statin therapy improves cardiovascular outcomes and has demonstrated plaque regression^44–46^ in multiple imaging studies, by the mechanism of modulating inflammation and decreasing or even depleting the lipid content within plaques^47–50^. However, the recent AIM-HIGH MRI sub-study^50^ showed that, despite two years of intensive lipid-lowering therapy, carotid arteries with IPH experienced larger increases in lipid core size and greater reductions in lumen area compared to plaques without hemorrhage, which exhibited stabilized plaque burden and significant reductions in lipid content. This suggests the IPH’s interplay with a potential non-lipid-driven pathway of plaque progression. One study^51^ identified CD163+ macrophages, induced by IPH, as potential contributors to a positive feedback loop involving increased vascular permeability, inflammation, and further IPH increase. Our findings similarly highlighted the role of IPH in counteracting the systemic stabilizing effects of contemporary medical treatment. While non-IPH plaques remained stable, current therapies may be insufficient to prevent IPH-associated plaque burden increase. Further investigation into the mechanisms behind this non-lipid-driven pathway is essential and could pave the way for novel therapeutic approaches specifically targeting IPH.

Overall, there remains a significant gap in longitudinal imaging studies investigating the pathophysiology of carotid atherosclerosis and IPH, as most studies are either short-term^7,9,52^ or involve long-term clinical follow-up without repeated imaging^23,39^. To the best of our knowledge, the longest imaging follow-up duration reported is 54 months^6^. However, this study was limited by a very small sample size and the use of 2D VWI, which provided inadequate coverage of the carotid bifurcation. This limitation was further compounded by challenges in identifying consistent imaging coverage for bilateral arteries and comparing multiple time points. Indeed, Geleri et al. reported that, when using the large-coverage 3D VWI sequence MERGE, only 54% of lesions were fully covered and 25% partially covered by 2D VWI in their analysis of 381 advanced plaques from the CARE-II study^53^. Recognizing these limitations, our study utilized MERGE, which provided adequate imaging coverage for bilateral and longitudinal analysis.

The emergence of deep learning-based imaging analysis also offers a promising solution to the labor-intensive nature of longitudinal VWI studies, reducing the need for extensive reviewer training and minimizing bias, as well-validated deep learning tools provide consistent and reproducible analyses across time points. While significant efforts have been made in segmenting the vessel wall on VWI^13–15^, robust algorithms for detailed plaque composition segmentation remain underdeveloped^54,55^. In our study, we successfully utilized deep learning to integrate multi-contrast VWI, demonstrating its potential to deliver more detailed and comprehensive assessments of plaque morphology and composition^56^.

Our study has a few limitations. First, although each patient underwent an average of five scans, the requirement to analyze the long-term evolution of atherosclerosis over more than five years restricted the number of participants in this retrospective analysis. Second, the retrospective nature of this analysis resulted in a relatively healthy and stabilized patient cohort, as individuals with severe symptoms—indicative of plaques experiencing dramatic reductions in stability— were more likely to drop out of the prospective cohort early. Additionally, the included subset had some differences in baseline characteristics compared to the full cohort^9^, including a higher proportion of males and lower prevalence of hypertension, which may further limit generalizability. Third, despite extensive follow-up, the absolute number of newly detected or resolved hemorrhages remained relatively low, underscoring the need for larger studies to better understand these potentially course-altering events in the plaque natural history. Fourth, our methodology did not perform spatial registration between MERGE and SNAP sequences or temporal registration across time points, relying instead on carotid bifurcation-based alignment to ensure spatial and temporal correspondence. While this landmark-based approach represents a practical solution as no established methodology exists for registration of 3D large-coverage multi-contrast VWI, residual misalignment may still exist, and precise tracking of identical anatomical locations over time remains limited. These remain important areas for future algorithm development. Finally, future research should aim to develop more advanced deep learning algorithms capable of simultaneously segmenting additional plaque components, such as lipid contents and calcification^57^, to further elucidate the complex interplay between these plaque components and the pathobiology of atherosclerosis.

## Conclusion

By leveraging deep learning-based segmentation on multi-contrast VWI, we successfully monitored long-term plaque dynamics across an average of five scans over a follow-up of six years in an asymptomatic cohort. Around half of the arteries with pre-existing IPH continued to exhibit it throughout the study period, while newly developed IPH also persisted, yet caused few new ischemic symptoms. Importantly, the presence of IPH was associated with disruption of the systemic symmetry of bilateral plaque progression: arteries without IPH demonstrated minimal growth under contemporary medical treatment, whereas the occurrence and volume of IPH were strongly associated with accelerated plaque burden progression.

## Data Availability

All data produced in the present study are available upon reasonable request to the authors.

## Abbreviations

IPH: intraplaque hemorrhage
VWI: vessel wall imaging
CEA: carotid endarterectomy
MERGE: motion-sensitized driven equilibrium prepared rapid gradient echo sequence
SNAP: simultaneous non-contrast angiography and intraplaque hemorrhage imaging
%WV: percent wall volume
%HV: percent hemorrhage volume

## Acknowledgement

This study was funded by National Institutes of Health under grant R01-HL103609, R01-NS125635 and R01-NS127317.

