## Supplementary Material for "Long-Term Carotid Plaque Progression and the Role of Intraplaque Hemorrhage: A Deep Learning-Based Analysis of Longitudinal Vessel Wall Imaging"

**Supplementary materials**

*Training datasets*

For the segmentation of carotid lumen and outer wall boundary using MERGE, the training data was provided by the 2020 and 2021 Grand Challenge hosted by the Society of Magnetic Resonance Angiography: the CARE-II dataset and the COSMOS dataset. The CARE-II dataset includes VWI data from 50 participants with expert annotations. The imaging was conducted on a 3T Philips MRI scanner using the MERGE sequence, producing isotropic voxels with a 0.35 mm resolution. Similarly, the COSMOS dataset offers VWI data from 50 participants with annotations. This data was collected using a 3T Philips MRI scanner with a 3D VISTA sequence, achieving isotropic voxels with a 0.35 mm resolution.

To segment IPH on SNAP, the training data were derived from the full study cohort of 98 subjects, using manually annotated IPH contours on baseline SNAP images^1^. These images, originally acquired with 0.4 mm isotropic resolution, were axially reformatted into 2mm slice spacing while maintaining the in-plane resolution. IPH was identified based on visual inspection, defined as hyperintense areas on SNAP in comparison to the sternocleidomastoid muscle.

*Validation datasets*

In a separate validation cohort, group A consisted of 14 subjects who were scheduled for carotid endarterectomy (CEA). Histological specimens were available and were matched with axially reformatted SNAP images, for assessment of IPH detection and segmentation^2^. Group B consisted of 33 patients who underwent baseline MRI (MERGE and SNAP) twice within one month. For each scan, the vessel wall and IPH were segmented from the MERGE and SNAP images, using the two abovementioned deep learning-based pipelines, for analysis of scan-rescan reproducibility. Detailed information on this validation cohort can be found in Figure 1 and the referenced publication^2^.

*Statistical analysis*

In group A, sensitivity and specificity for IPH detection were estimated at the slice level, and agreement in IPH detection between deep learning and histological analysis was evaluated using Cohen’s kappa. Agreement in IPH area quantification between SNAP and histology was assessed using Spearman’s correlation coefficient, as it is less affected by shrinkage of histological specimen during processing compared to absolute agreement measures like the intraclass correlation coefficient (ICC) and the Bland-Altman plot.

In group B, Cohen’s kappa was used for evaluating the scan-rescan reproducibility of IPH detection of the deep learning-based image segmentation pipeline, and the ICC was used for evaluating the reproducibility of plaque burden and IPH volume (within arteries identified as IPH+ in either scan). The nonparametric bootstrap and percentile method was used to calculate 95% confidence intervals (CI).

*Assessment results of deep learning-based plaque segmentation*

In the 14 arteries in group A, a total of 121 SNAP slices were matched with histological specimens, of which 49 (40%) slices from 13 (93%) plaques had histology-detected IPH. Deep learning-based analysis had moderate overall agreement with histology in detecting IPH (kappa: 0.62, 95% CI: 0.48 - 0.76). The sensitivity and specificity for IPH detection at the slice level were 61% and 97%, respectively. In these 30 slices where both methods agreed on IPH presence, IPH area measured using deep learning (mean ± SD: 16.4 ± 12.6 mm^2^) and histology (mean ± SD: 12.0 ± 7.3 mm^2^) were strongly correlated (Spearman’s rho = 0.63, p < 0.001).

In the 33 subjects with 66 arteries in validation group B, eight pairs of MERGE images were excluded due to low image quality on either scan, and one artery was lesion-free. In the remaining 57 pairs of arteries, %WV characterized by MERGE showed high scan-rescan reproducibility, with an ICC of 0.88 (95% CI: 0.80 - 0.93). Mean lumen area and mean total vessel area also demonstrated excellent reproducibility, with ICCs of 0.97 (95% CI: 0.95 - 0.98) and 0.95 (95% CI: 0.92 - 0.97), respectively.

In SNAP from group B, 14 (24.6%) arteries were categorized as IPH+ in both scans, 3 (5.3%) were categorized as IPH+ either scan, and 40 (70.2%) were categorized as IPH- in both scans, yielding a kappa of 0.87 (95% CI: 0.71 - 0.99). Of the 17 arteries categorized as IPH+ in at least one scan, IPH volume showed high reproducibility, demonstrated by an ICC of 0.94 (95% CI: 0.84 - 0.98).

Bland-Altman plots indicated no apparent relationship between variance and mean for measurements of %WV, mean lumen/total vessel area and IPH volume (see Supplementary Figure 2).

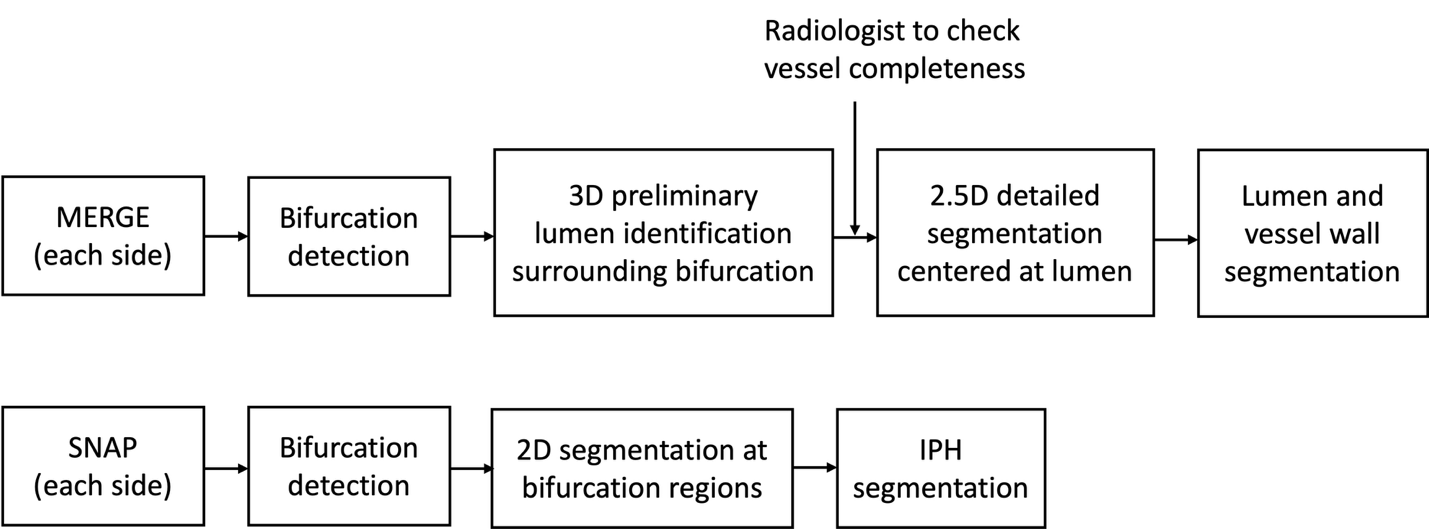

Supplementary Figure 1. Deep learning pipeline flow chart showing the processing workflows for MERGE and SNAP sequences. Each side (left and right carotid) is processed independently through the respective pathways.

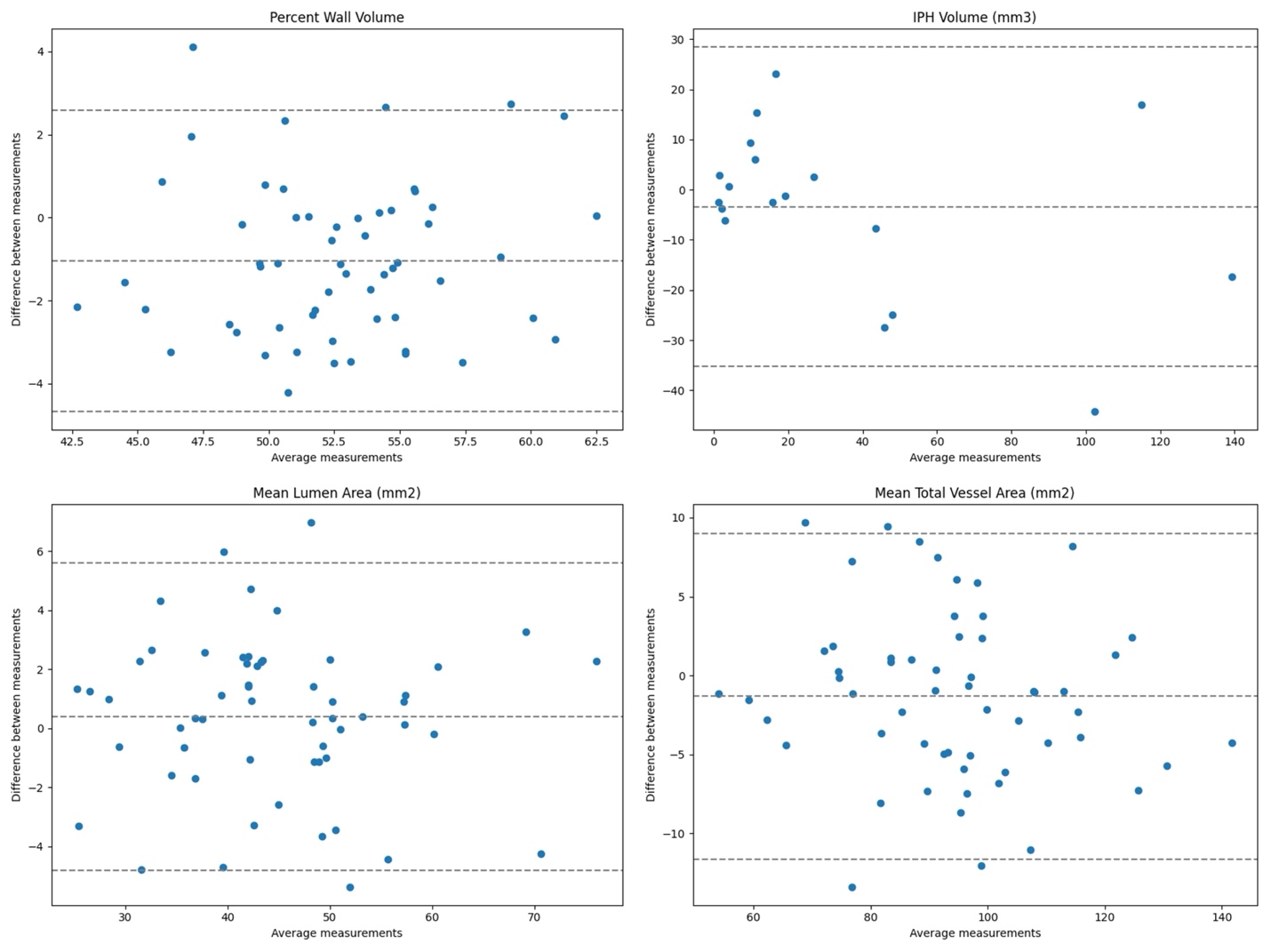

Supplementary Figure 2. Bland-Altman plots demonstrate scan-rescan reproducibility for deep learning-based measurements of percent wall volume, IPH volume, mean lumen area and mean total vessel area.
